# Carbapenemase genes in hospital wastewater in Africa: A systematic review and meta-analysis

**DOI:** 10.1101/2025.03.26.25324662

**Authors:** Oumou Hamidou, Abdourahamane Yacouba, Ounoussa Tapha, Harouna Moussa, Ismael Illa Salifou, Souleymane Brah, Saidou Mamadou, Lamine Said Baba-Moussa

## Abstract

Hospitals may be a significant reservoir of antibiotic-resistant genes, including carbapenemase genes. This study aimed to evaluate the prevalence and distribution of carbapenemase genes in hospital wastewater in Africa.

We conducted a comprehensive search on carbapenemase genes in hospital wastewater in Africa up to November 31, 2024, using PubMed, Google Scholar, and African Journal Online (AJOL) databases. We included original studies without time or language restrictions. The meta-analysis used the R package ‘metafor’ with a random effects model due to expected heterogeneity among studies. Heterogeneity was assessed using the I^2^ statistic.

We included 13 studies conducted in seven African countries. Compared with those in treated wastewater, carbapenem genes were more enriched in raw wastewater. Among the 13 different carbapenemase genes reported, 9 (69.2%) genes were specific to the raw wastewater group, including *blaIMP*, *blaVIM, blaOXA-181, blaOXA-69, blaOXA-1-like, blaOXA-48-like, blaGES, blaOXA-416,* and *blaOXA-51*. They are more commonly isolated from enterobacteria (7; 58.3%). The estimated overall prevalence was 23.8% (95% CI: 15.4% 1–33.4%), and the heterogeneity between studies was substantial (I^2^ = 96.1%; p<0.01).

This review emphasizes the presence of carbapenemase genes in hospital wastewater in Africa, showing a high prevalence. African hospitals should focus on controlling the spread of these genes in wastewater and prioritize the safety of health professionals and patients.

**IMPORTANCE:** Antibiotic resistance poses a significant threat to global public health. It is a complex process influenced by host, environmental, and pathogen factors. Effluents, especially from hospitals, are substantial sources of antibiotic-resistant bacteria due to high bacterial loads, nutrients, and low levels of antibiotics. Improper handling and disposal of hospital effluents can pose a risk to public health by promoting the spread of resistance genes. Sanitation and hospital waste management standards in Africa vary significantly. Hospital wastewater can be a significant source of antibiotic-resistance genes, including carbapenemase genes. Carbapenemases can hydrolyze penicillins, cephalosporins, and carbapenems, making them a serious public health concern as carbapenems are often the last resort antibiotics for treating multiresistant bacteria. This review aims to assess the prevalence and distribution of carbapenemase genes in hospital wastewater across Africa.

## INTRODUCTION

The emergence and spread of antibiotic resistance represent a real threat to global public health (1). The emergence of antibiotic resistance is a complex process that often involves host, environmental, and pathogen factors (2). Studies have shown that aquatic environments represent one of our planet’s most important microbial habitats. Aquatic environments act as reservoirs through which microorganisms, particularly those with high levels of antibiotic resistance, are widely disseminated among the natural environment, humans, and animals (3, 4).

However, effluents can be considered important hotspots for the presence and potential proliferation of antibiotic-resistant bacteria (5). In addition to high bacterial loads and a nutrient-rich environment, hospital effluents in particular can contain subtherapeutic concentrations of antibiotic agents, which can facilitate the emergence and spread of resistance genes from one bacterium to another (6). These hospital effluents can therefore compromise public health if handled and disposed of incorrectly (7, 8).

Additionally, the resulting compounds and biological associations in effluents can spread into the environment and ultimately affect human and animal populations (9). Hospital wastewater is a major source of antibiotics and antibiotic-resistant bacteria (ARB) (10), and this threat is not known in sub-Saharan aquatic environments (11). Hospital wastewater can be a major reservoir of antibiotic-resistance genes, including carbapenemase genes. Carbapenemases can hydrolyze penicillins, cephalosporins, and carbapenems, limiting antibiotic treatment options (12). The emergence of these enzymes has increasingly been described worldwide and represents a real public health problem, as carbapenems are very often the last active molecules in the therapeutic arsenal available to combat multiresistant bacteria.

In Africa, sanitation and hospital waste management standards can vary considerably, and hospital wastewater can constitute a major reservoir of antibiotic-resistance genes, including carbapenemase genes. Thus, this meta-analysis study aimed to determine the prevalence and distribution of carbapenemase genes in hospital wastewater in Africa.

## METHODS

### Study design

Relevant studies that have reported on carbapenemase resistance genes in bacteria isolated from hospital wastewater in Africa were systematically searched. A protocol for this review has been registered with the International Prospective Register of Systematic Reviews (PROSPERO) (registration number: CRD42024526818).

### Search strategy and selection criteria

A systematic literature search was conducted to identify articles reporting on the prevalence of carbapenemase genes in hospital wastewater in Africa. Several searches were carried out in PubMed, Google Scholar, and African Journal Online (AJOL). Our search strategy used various relevant keywords: “prevalence” AND (“carbapenemase genes” OR “blaKPC, blaNDM, and blaOXA-48-like genes”) AND “wastewater ‘AND ‘hospitals” AND (“Africa”, OR “names of 54 African countries”). This study was designed according to the PRISMA (Preferred Reporting Items for Systematic Reviews and Meta-Analyses) guidelines (13).

The identified publications were reviewed up to November 30, 2024. The searches were not restricted by language. Two authors independently performed the literature search. After removing duplicates, studies identified in the initial search were first screened by title and abstract and retained if they met the following predefined inclusion criteria, as follows: (i) original article published or accepted in a peer-reviewed journal, (ii) studies investigating the prevalence of carbapenemase genes in hospital wastewater samples, and (iii) studies conducted in one of the 54 African countries,(iv) studies using molecular detection methods (PCR, sequencing) and immunochromatographic testing to identify carbapenemase genes.

The full texts of the articles meeting the inclusion criteria were retrieved and examined in greater detail. Journal studies (narrative reviews, systematic reviews, and meta-analyses), letters to the editor, and opinions were excluded. Additionally, studies conducted outside the specified period of this systematic review, studies involving sample types other than hospital wastewater, and studies using detection methods other than PCR, sequencing, or immunochromatographic testing were excluded. The selection process is documented in the PRISMA flowchart of study selection (13). Three reviewers (HO, TO, YA) independently screened the titles and abstracts of the studies and assessed the potential eligibility of the relevant full texts. Reviewers resolved any discrepancies in their selections through discussion.

### Quality assessment

Independent reviewers (TO, YA) critically analyzed the included studies to ensure that the findings were reliable and consistent. The mixed methods appraisal tool (MMAT)(14), a critical appraisal tool, was used to assess the quality of the included studies.

### Data extraction process

Two reviewers (HO, TO) independently extracted appropriate data from each eligible study, resolving disagreements through discussion. A third researcher (YA) confirmed the eligibility of the included studies before they were included in the analysis.

The following variables were extracted: first author’s name, year of publication, country, study type, number of samples, sample type, prevalence of carbapenemase resistance, technique used, bacterial species isolated, resistance phenotypes, sequence type, resistance genotypes, antibiotic resistance, resistance type, and resistance plasmids.

### Data synthesis and analysis

The data were analyzed via R version 4.4.1. The prevalence and distribution of carbapenemase genes were grouped by country, hospital type, and detection method. A narrative synthesis of the results was carried out, highlighting trends, geographical variations, and factors associated with carbapenemase gene prevalence. For the meta-analysis, the packages meta (15) and metafor (16) were used to calculate the pooled percentage and 95% confidence interval (CI) using a random-effects model. Subgroup meta-analyses were performed according to the prevalence of carbapenemase genes by bacteria group and by sample source. Statistical heterogeneity between studies was evaluated for the significance of heterogeneity and the inconsistency index, where I^2^ > 75% and a significance level < 0.05 (p-value) were deemed to represent substantial heterogeneity. Only studies that reported the proportion of carbapenemase genes detected in raw and/or treated wastewater were considered for meta-analysis. A funnel plot and Egger weighted regression test were performed to detect the presence of publication bias in the meta-analysis. The asymmetry of the funnel plot and significant (p-value < 0.05) Egger tests indicated significant publication bias.

## RESULTS

A total of 982 relevant and unduplicated studies were identified and extracted from the databases. After the titles and abstracts were examined, 965 articles were deemed irrelevant and excluded. Of the 17 eligible full-text articles of interest, only 13 met the inclusion criteria and were included in the final analysis (Figure 1). On the basis of the included studies, the first article on the prevalence and distribution of carbapenemases in hospital wastewater in Africa was published in 2017.

**Figure 1:**
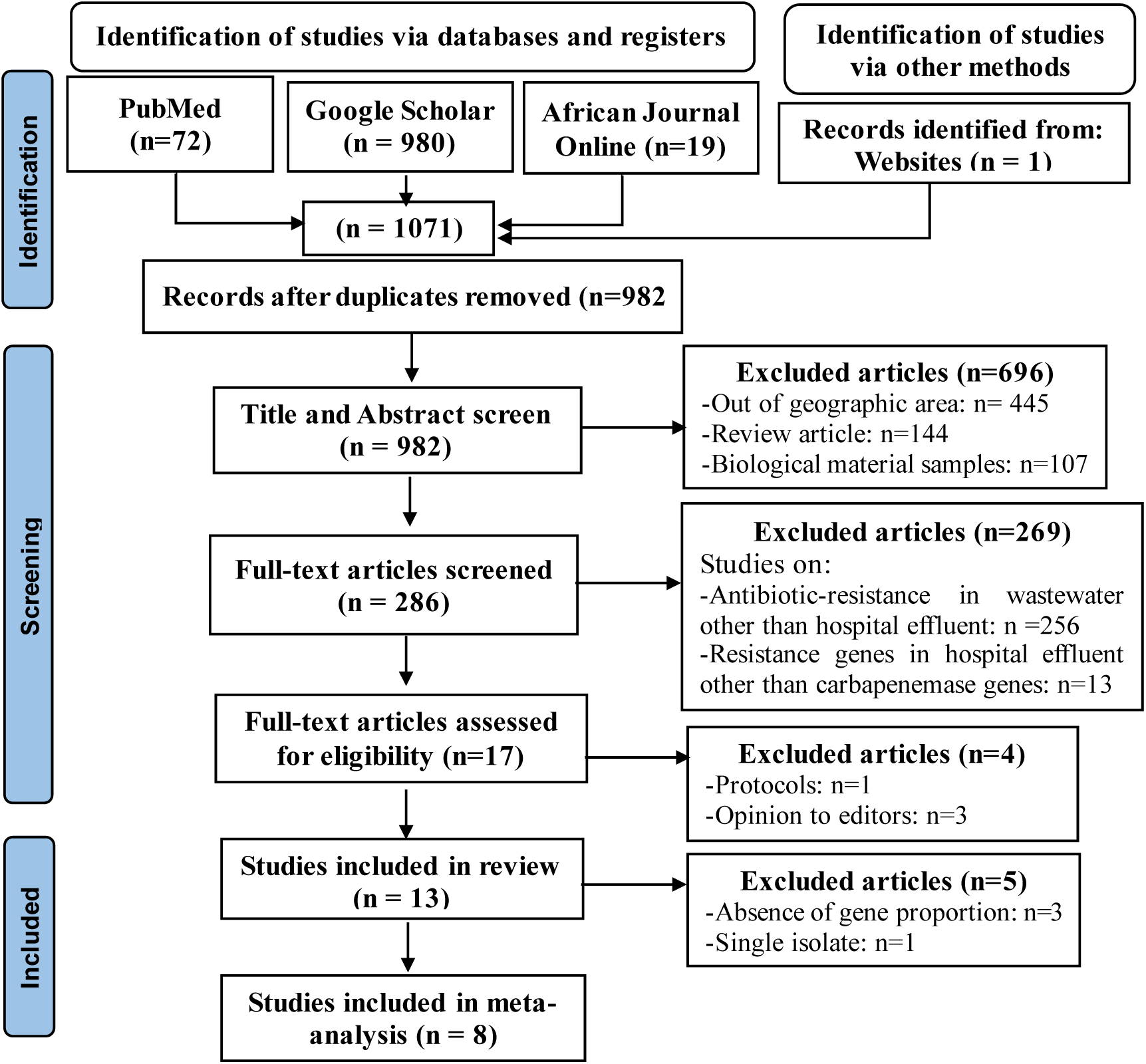
PRISMA flow diagram of study selection.

All 13 studies were cross-sectional (Table 1). Raw wastewater was collected as the sample type in all included studies, except for three studies (17–19), where treated wastewater was used as the sample type. Disc diffusion was the common method used for antimicrobial susceptibility testing, except for two studies where the broth microdilution method (18) and VITEK®2 (19) were used. PCR or PCR sequencing was used to detect the carbapenemase genes, except in one study where the immunochromatographic test was O.K.N.V.I. RESIST-5 (CORIS BioConcept, Belgium) (20) was used for the detection of carbapenemase genes.

**Table 1:**
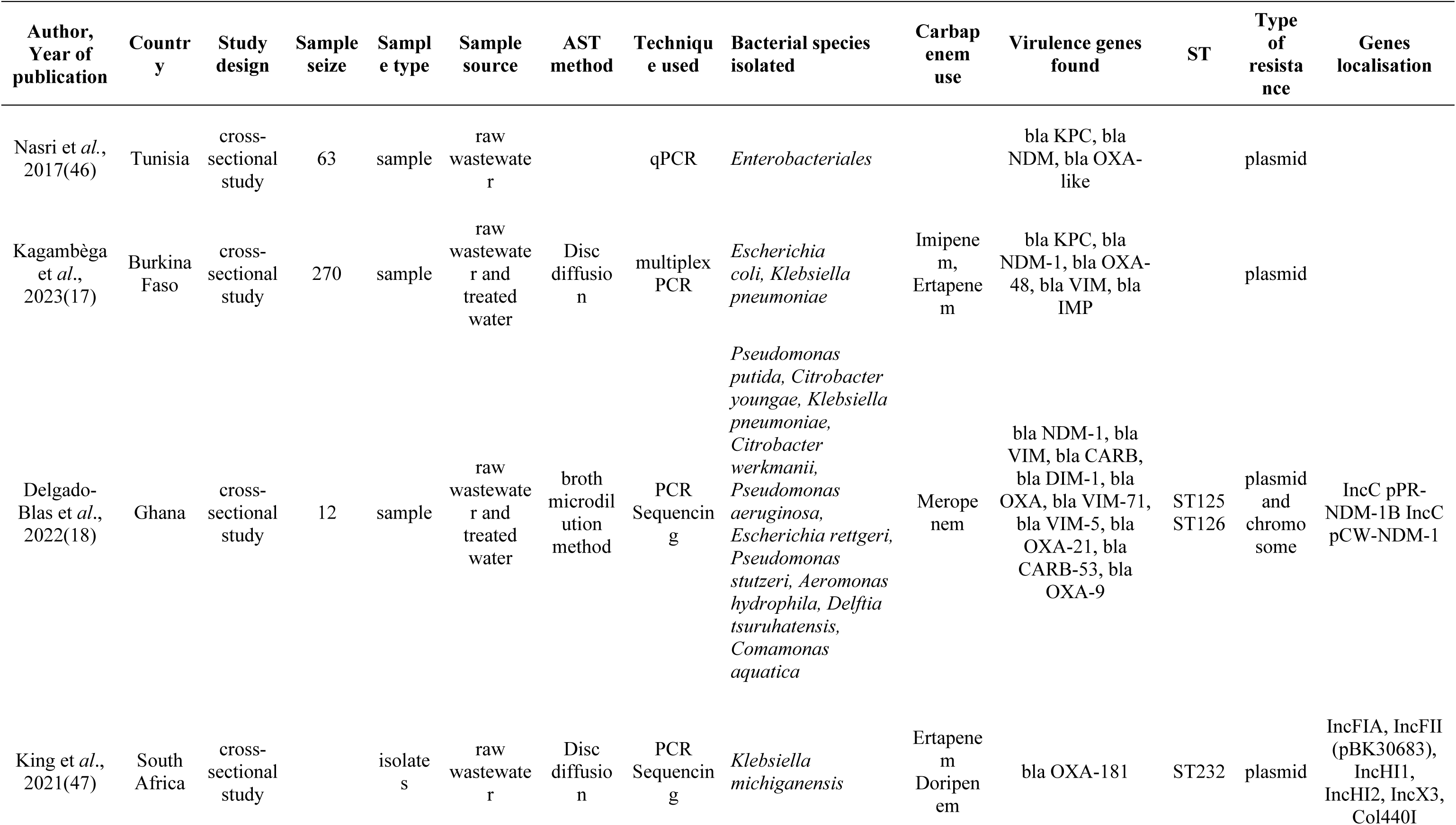
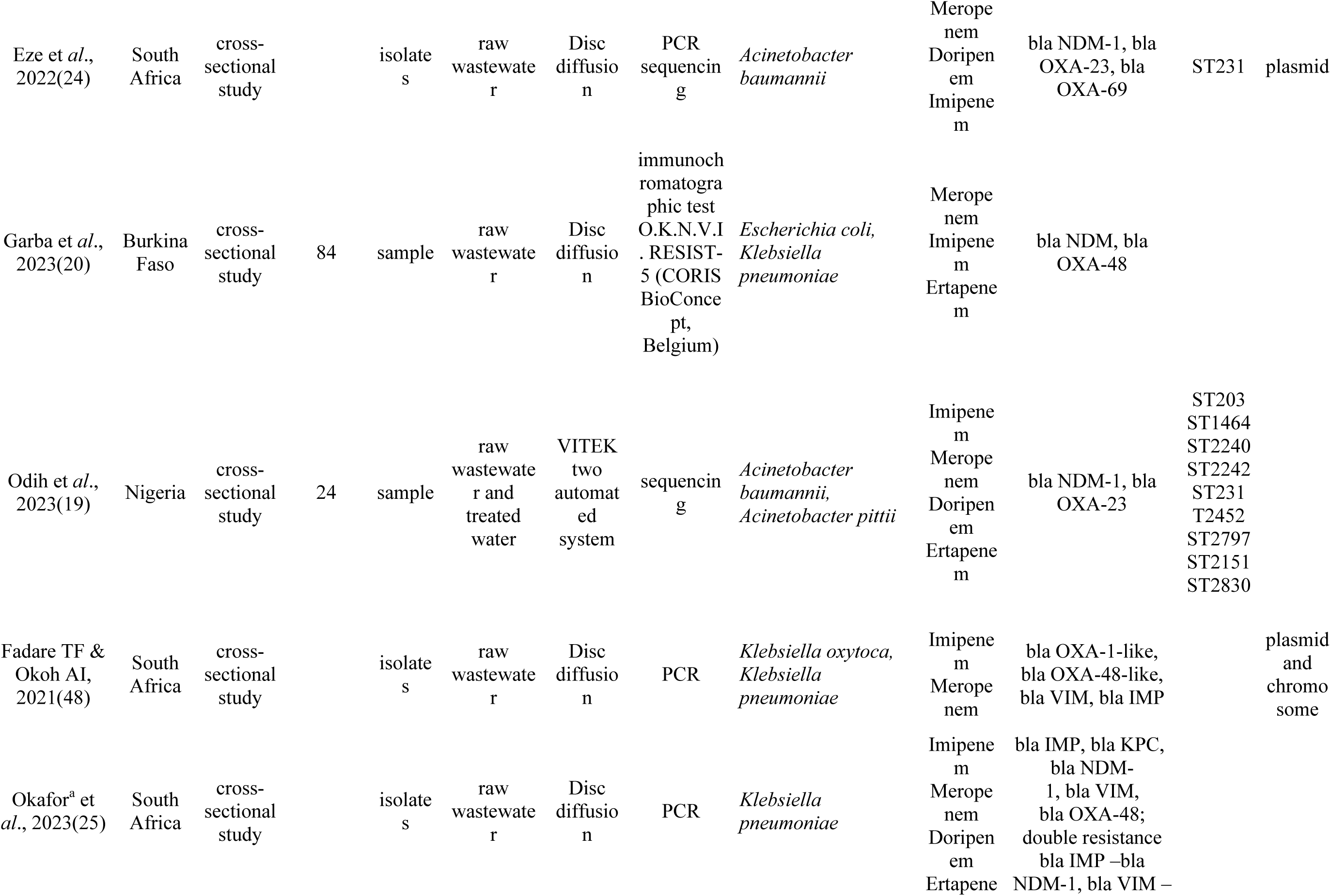
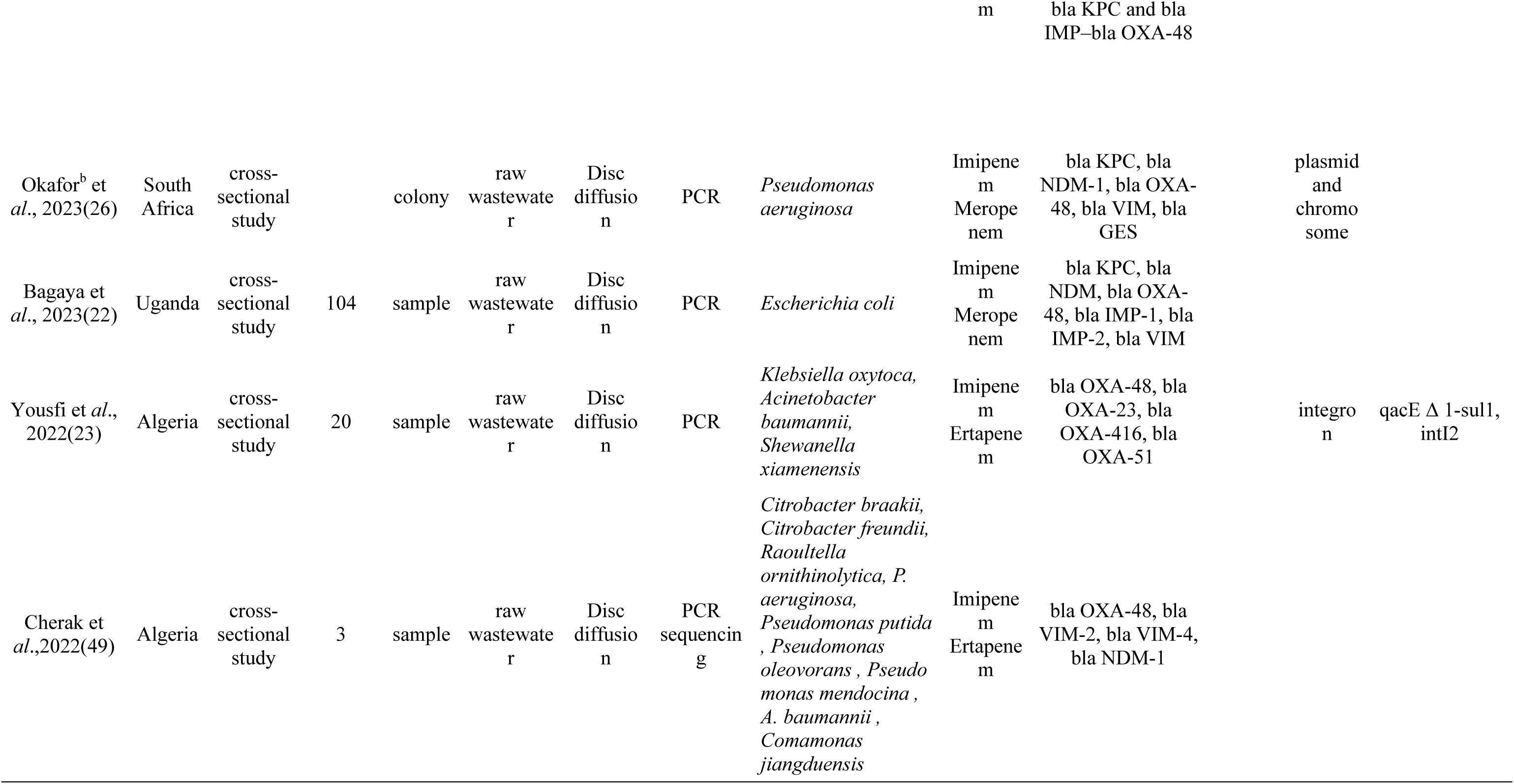
Characteristics of eligible research papers.

### Characteristics and distribution of carbapenemase genes in Africa

This review involved 13 studies conducted in seven countries: South Africa (n=5), Burkina Faso (n=2), Tunisia (n=1), Uganda (n=1), Nigeria (n=1), Ghana (n=1), and Algeria (n=2) (Figure 2). The studies were published between 2017 and 2023. The studies revealed carbapenemase resistance genes in bacteria isolated from hospital untreated wastewater (8; 72.7%) and untreated and treated wastewater (3; 27.3%). The Venn diagram (Figure 3) demonstrates the shared and exclusive carbapenemase genes between the treated and raw wastewater. The total number of different carbapenemase genes detected was 13, among which 4(30.8%) genes were shared by both groups, and 9 (69.2%) genes were specific to the raw wastewater group, indicating a decrease in the carbapenemase gene diversity as the hospital wastewater was treated.

**Figure 2:**
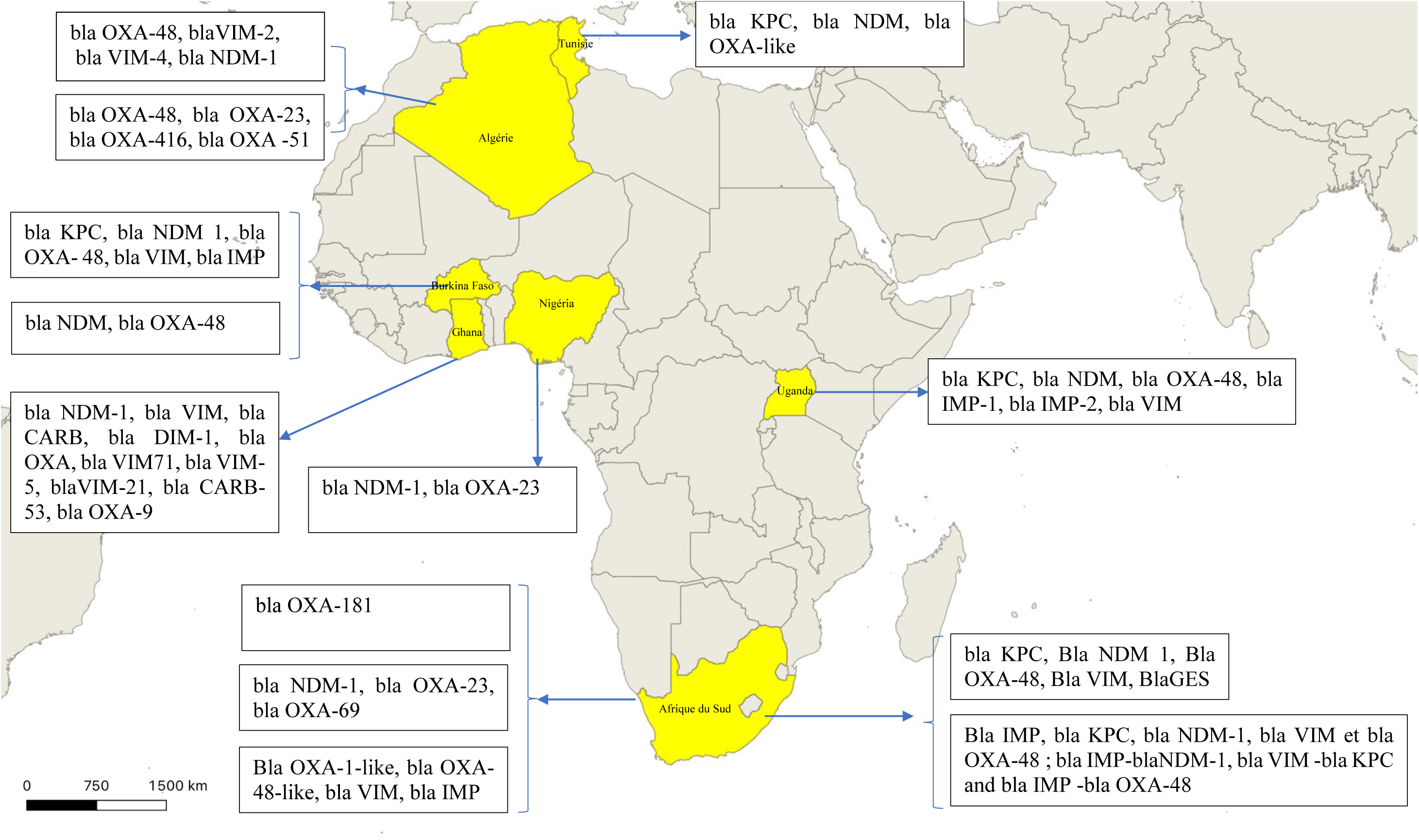
Geographical distribution of carbapenemase genes in Africa.

**Figure 3:**
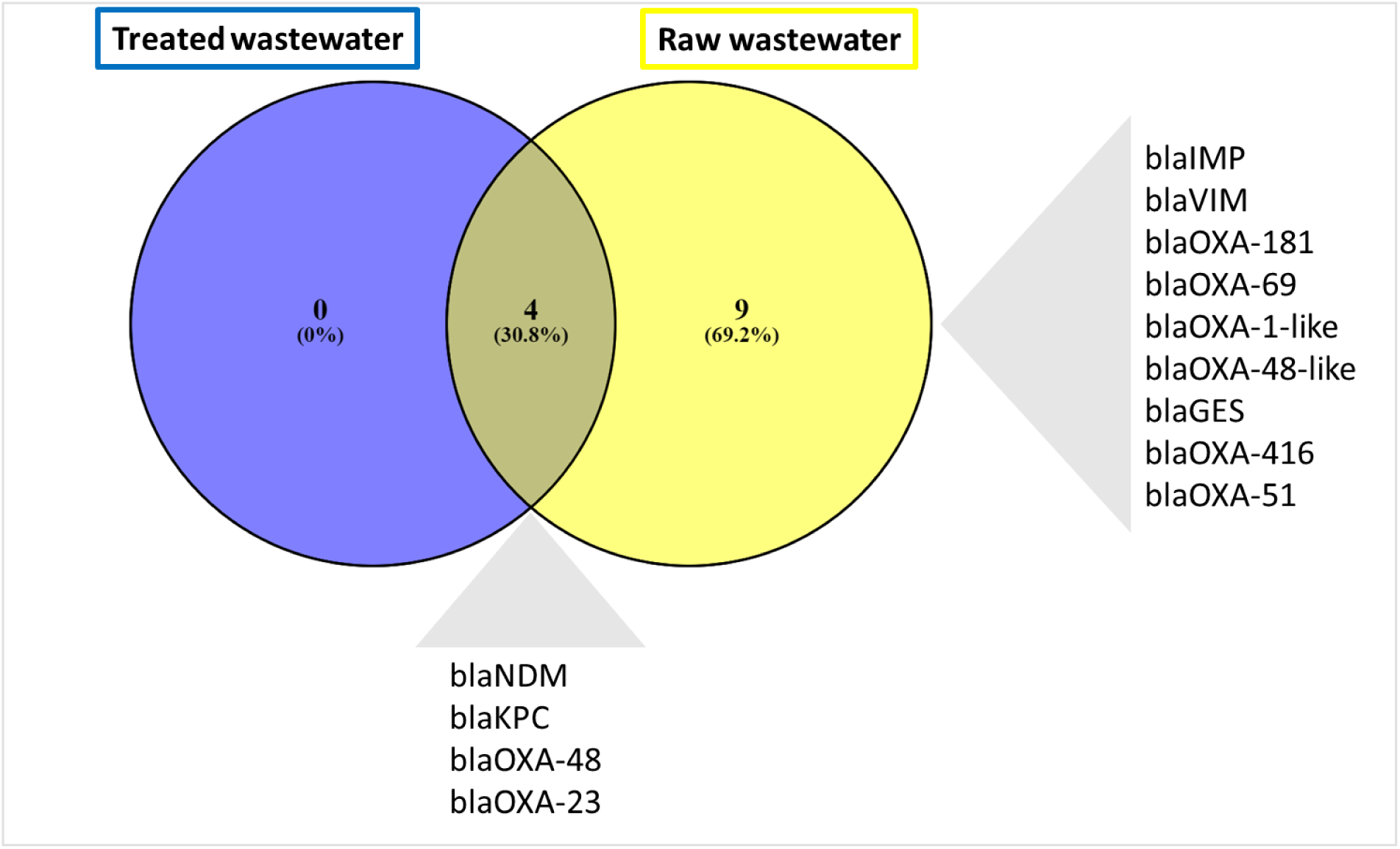
Venn diagram distribution of carbapenemase genes between treated and raw hospital wastewater.

### Bacteria harboring carbapenemase genes

Carbapenemase genes were described mainly in enterobacterial species (7; 58.3%), non-fermentative bacteria (3; 25.0%), and enterobacterial and non-fermentative bacteria (2; 16.7%). At the species level, carbapenemase genes were most frequently observed in *Klebsiella pneumoniae* (19,23%), followed by *Escherichia coli* (11,53%) and *Acinetobacter baumannii* (15,38%).

### Mechanism of resistance

Chromosomal and plasmid-mediated mechanisms have been reported, with plasmid-mediated carbapenemase resistance genes (6; 85.7%) most frequently reported.

### Techniques used to detect carbapenemase

Four studies used PCR alone or PCR sequencing to detect the carbapenemase genes, whereas only one used an immunochromatographic test.

### Pooled prevalence estimates of carbapenemase genes in Africa

As shown in Figure 2, of the 13 articles included, 10 (8, 17, 19–26) provided data on the proportion of carbapénèmases genes. Among the 10 studies, two (8, 24) involved a single isolate and were not included in the meta-analysis. The pooled prevalence of carbapenemase genes in Africa was 23.8% (95% CI: 15.4% 1–33.4%). Significant heterogeneity was observed according to the I^2^ test (p<0.01) (Figure 4).

**Figure 4:**
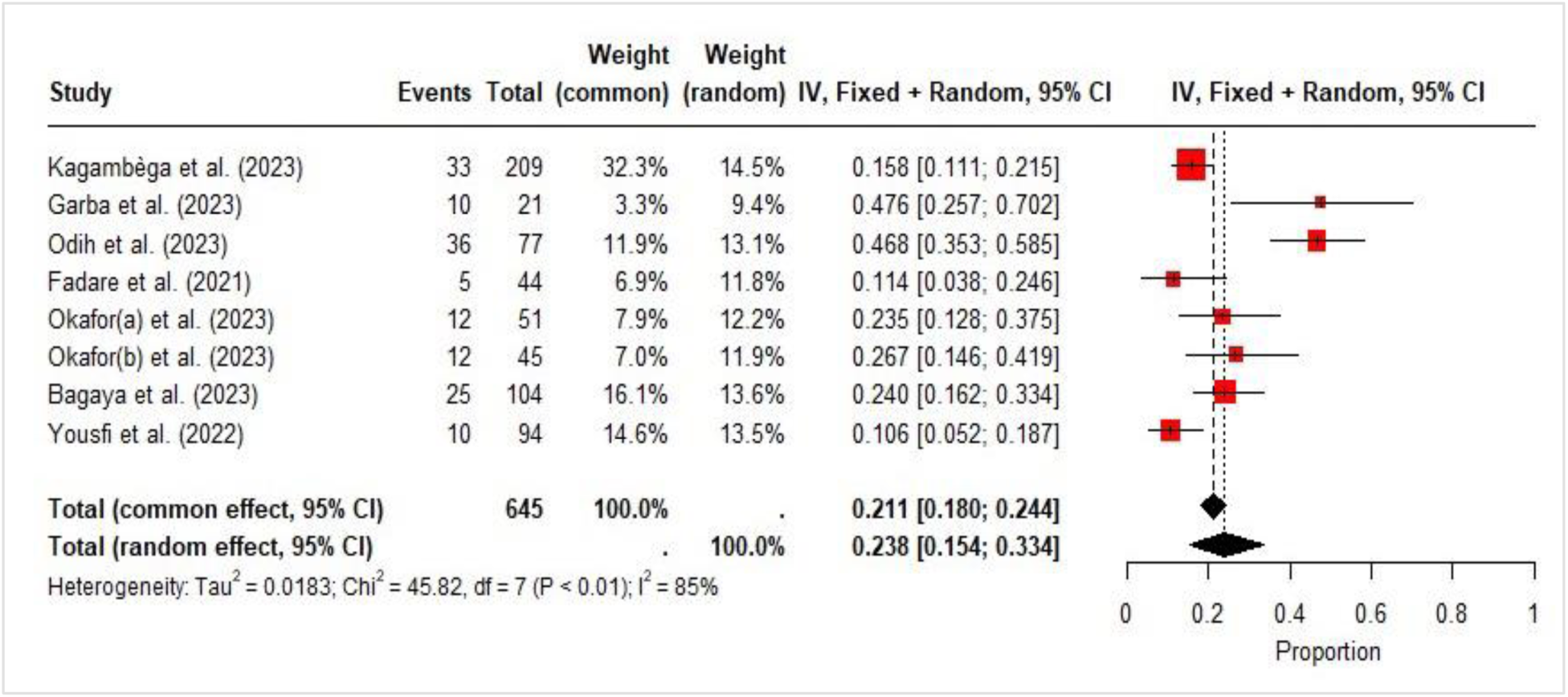
Prevalence of carbapenemase genes in wastewater.

### Subgroup analysis

The information in Figures 5 and 6 outlines the subgroup analysis of the prevalence of carbapenemase genes by bacterial groups and by sample source. Subgroup analysis by bacterial group revealed that the non-fermentative bacterial group (37%; 95% CI: 17.0– 57.0%) had a greater proportion of carbapenemase genes than did the enterobacterial group 21%; 95% CI: 13–30%) (Figure 5). Subgroup analysis by sample source revealed higher rates of carbapenemase genes in the raw and treated wastewater (31%; 95% CI: 1 – 61%) than in the raw wastewater (21%; 95% CI: 13-30%) (Figure 6).

**Figure 5:**
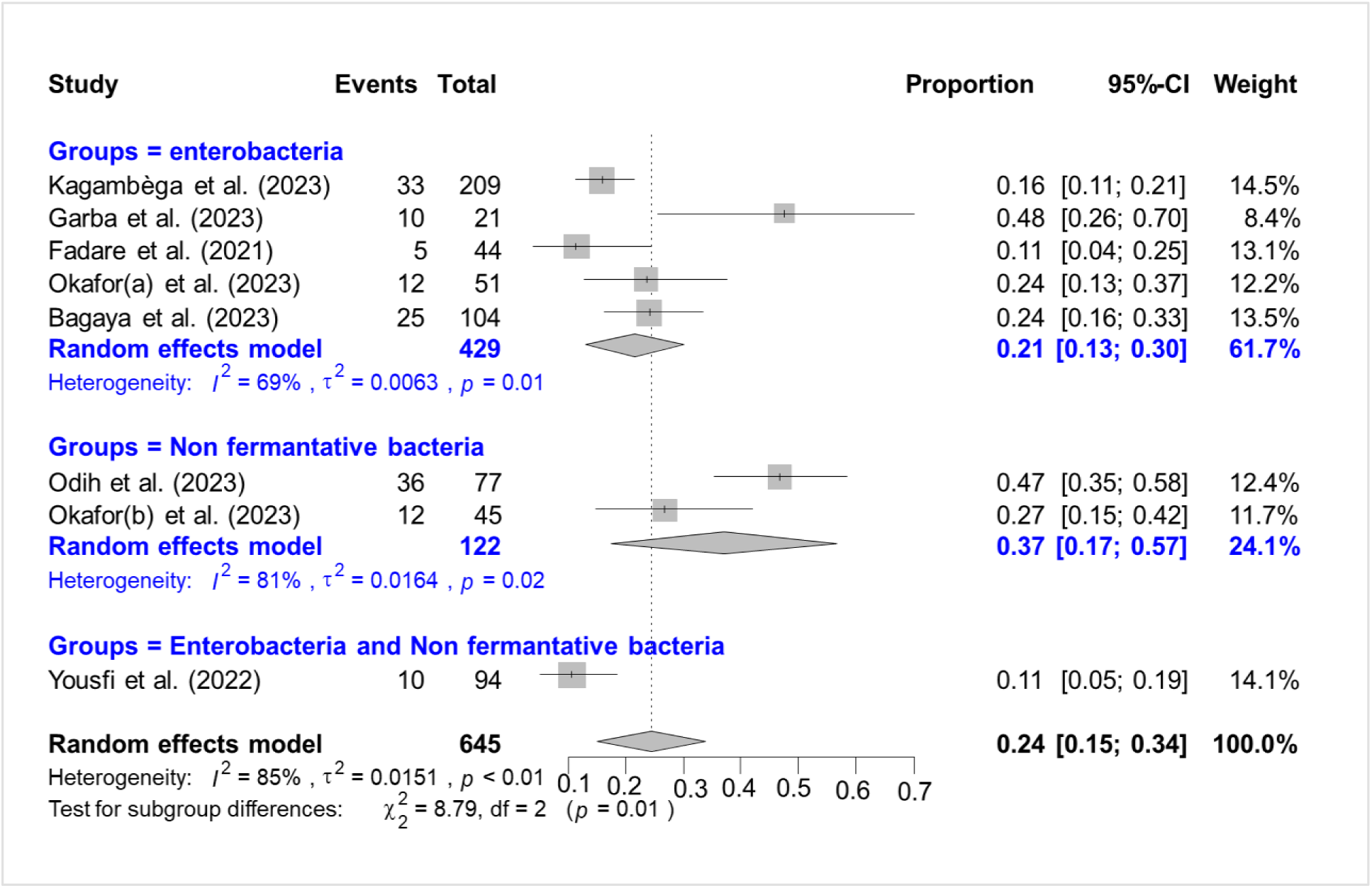
Subgroup analysis by bacterial groups.

**Figure 6:**
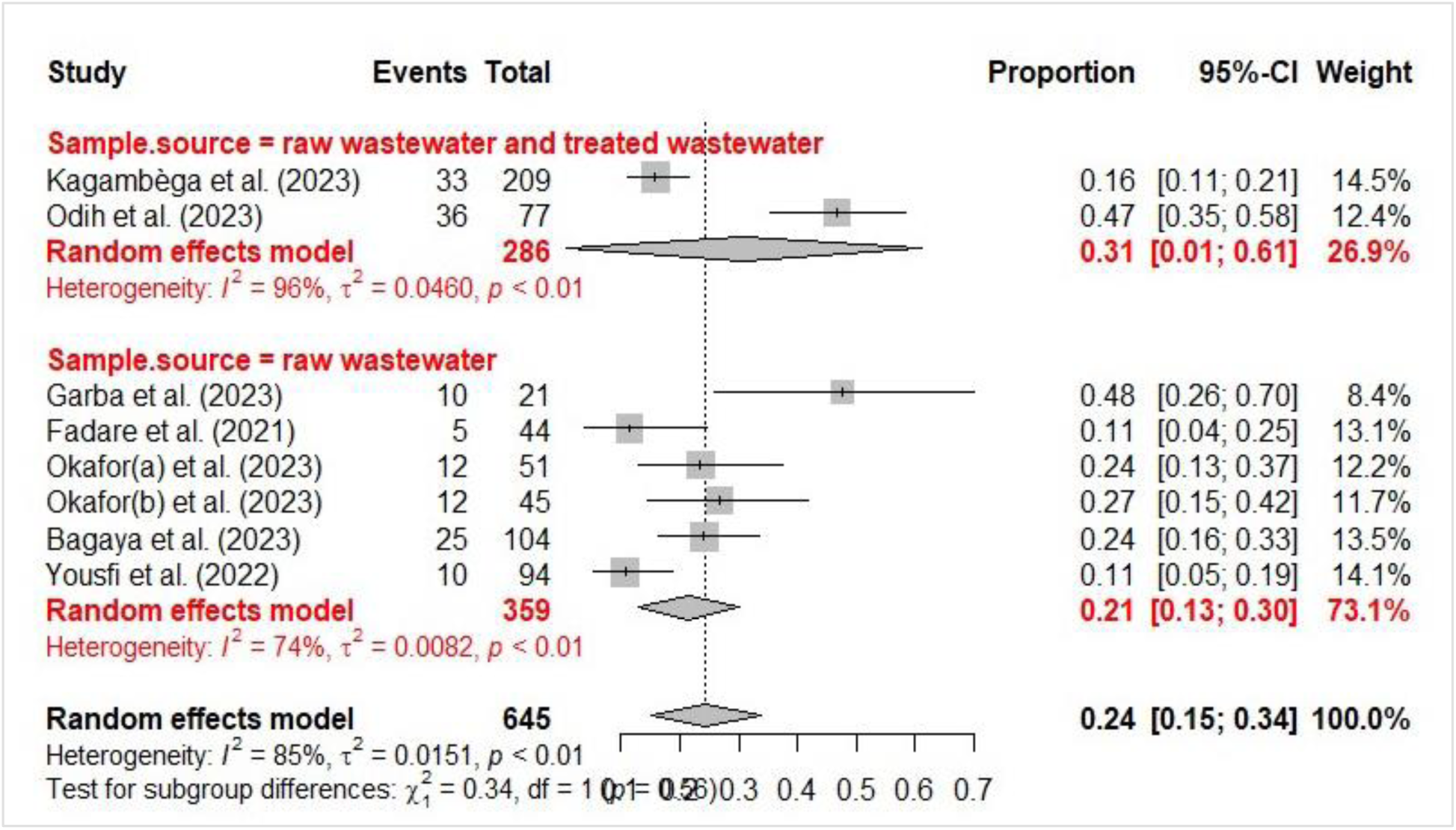
Subgroup analysis by sample source.

### Publication bias

The studies included were evaluated visually for possible publication bias via a funnel plot (Supplementary Figure 1). The funnel plot asymmetry indicated publication bias, as 62.5% of the studies fell on the right side of the triangular region (Supplementary Figure 1). However, the results of Egger’s test revealed no significant publication bias (p = 0.273)

## DISCUSSION

Carbapenems are broad-spectrum antimicrobials of last resort. Resistance to carbapenems is a public health threat, leading to an increase in infection mortality, hospital stay duration, and cost of treatment (27). Resistance to these antibiotics, which is mainly mediated by enzymes such as carbapenemases (KPC, NDM, OXA-48), poses a global public health challenge (27–29). Hospital wastewater is a key environment for the dissemination of these resistance genes because of its high degree of contamination with pathogenic bacteria, antibiotics, and biological residues (30).

This systematic review and meta-analysis were based on 13 articles that focused on the antibiotic resistance and carbapenemase resistance genes of bacteria isolated from hospital wastewater. The estimated overall prevalence of carbapenemase resistance genes was 23.8% (95% CI: 15.4% 1–33.4%), and the heterogeneity between studies was substantial (I2 = 96.1%; p<0.01). Studies have revealed an alarming prevalence of carbapenemase genes in hospital effluent worldwide (17, 28, 31, 32), particularly in regions where wastewater treatment infrastructures are inadequate, as in many African countries. These results illustrate the importance of antimicrobial resistance on continents. This high estimate reflects the impact of medical practices, wastewater management systems, and poor infrastructure resources in controlling resistant bacteria.

The Venn diagram shows a reduction in carbapenemase genes after the treatment of raw wastewater. Other studies have also shown a reduction in carbapenemase genes in wastewater after treatment, confirming the partial effectiveness of current treatment systems. Studies conducted in Brazil and China(33, 34) on a hospital wastewater treatment system have also shown that hospital wastewater treatment may not be fully effective in eliminating multidrug-resistant bacteria and resistance genes from hospital wastewater (35).

Zhang et al 2020 in China reported carbapenemase genes in treated wastewater in China (36). This could be explained by the fact that hospital effluents contain large quantities of antibiotic and antiseptic residues, which could favor the development of antimicrobial-resistant bacteria. these compounds exert a selective pressure favoring antimicrobial-resistant bacteria even when they are present in very low concentrations. It has been suggested that nutrient-rich environments offer optimal conditions for horizontal gene transfer, including the passage of plasmids and transposons encoding antibiotic resistance (37) and the transformation of nonpathogenic bacteria into resistance gene reservoirs (30).

However, work by Cahill et al. highlights that carbapenemase-producing Enterobacteriaceae (EPC) are frequently found in hospital effluent, both upstream (prehospital) and downstream (posthospital) of treatment (28). This can be attributed to the ability of treatment plants to eliminate or reduce resistant bacteria and genes through physicochemical and biological processes. However, this reduction does not imply complete elimination, suggesting that conventional treatment systems may be insufficient to fully manage microbiological contaminants. Particularly in Africa, where treatment systems are often rudimentary or poorly maintained, they are clearly insufficient to eliminate these microbiological contaminants. As shown by Hounmanou et al., hospital wastewater treatment is not commonplace in many low-income countries in Africa (38). In countries where wastewater treatment is in place, it generally involves a combination of physical, chemical, and biological processes to remove solids and organic matter. However, these systems often prioritize the removal of conventional pollutants, and are generally insufficient to eliminate antimicrobial-resistant microorganisms and antibiotic-resistance genes.

The persistent presence of resistance genes after treatment highlights the need for advanced treatment technologies, such as ozonation, ultrafiltration, or the application of specific bioprocesses to eliminate resistant genes (39).

This study revealed a high diversity of carbapenemase genes from all Ambler classes in African wastewater, although not all regions were represented. Notably, strains containing several genes encoding carbapenemases have been identified (40).

These genes can be transferred horizontally between different bacterial species via mobile genetic elements such as plasmids. (41) reported evidence of possible environmental transfer of the bla KPC plasmid between strains of *K. pneumoniae*, and Betteridge et al reported that there is likely to be an environmental exchange of intergenic plasmids between Enterobacteriaceae (42). Resistance genes and associated mobile genetic elements, therefore, need to be characterized in outbreak investigations of carbapenem-resistant bacteria(41).

To date, data on carbapenemase genes in Africa are scarce. Prospective longitudinal studies using a One Health approach and simultaneously assessing carbapenem resistance genes in humans, animals, and the environment are needed to better understand the main factors involved in the spread of carbapenemase genes in Africa (43).

Carbapenemase genes have been described mainly in *Enterobacteriaceae* species (7; 58.3%), nonfermentative bacteria (3; 25.0%), and *Enterobacteriaceae* and nonfermentative bacteria (2; 16.7%). In terms of species, carbapenemase genes were most frequently observed in *Klebsiella pneumoniae*, followed by *Escherichia coli* and *Acinetobacter baumannii*. These results confirm the crucial role of nonfermentative bacteria as major reservoirs of resistance, particularly in hospital environments where these opportunistic bacteria are ubiquitous. These nonfermentative bacteria are often associated with serious nosocomial infections that are difficult to treat. These findings may reflect biological differences in which *Pseudomonas* spp. and *Acinetobacter* spp. are environmental colonizers that survive under low nutrient conditions, whereas *Enterobacteriaceae* are predominantly of human origin, and their concentrations in sewage may represent different ecosystems and inoculations directly from patient waste. However, all of these organisms are capable of colonizing biofilms in the water system (44).

A high degree of heterogeneity in overall prevalence was also observed in a systematic review of the prevalence of carbapenemase-producing *Enterobacteriaceae* in clinical samples in Ethiopia by Alemayehu et al.(45); this significant heterogeneity highlights the marked differences between the included studies, probably due to methodological variations, geographical differences, or socioeconomic contexts. This highlights the importance of harmonized studies in the future, with standardized methodologies to better assess the prevalence of resistance genes.

However, our study has certain limitations. First, a funnel plot with a small number of fewer than 10 studies may appear asymmetric simply owing to chance. Indeed, the power of the tests is too low to distinguish chance from real asymmetry. Second, the representativeness of individual studies within countries is unclear in most cases, and third, all the studies included in this systematic review were cross-sectional studies. The studies were not evenly distributed across Africa. Some countries have more studies included in the analysis, whereas others have none. This meta-analysis has several limitations, such as the presence of significant heterogeneity even after subgroup analysis for some variables. These limitations may have affected the results reported in this review concerning the overall prevalence of carbapenem-resistance genes in wastewater.

## CONCLUSION

This study reported on the prevalence of carbapenemase genes in hospital wastewater in Africa. Although data from Africa are still limited, this systematic review provided evidence that carbapenemase genes in hospital wastewater are widespread in Africa. Further studies are needed to identify the links between the release of carbapenem resistance genes from hospital wastewater treatment plants, their presence in the surrounding environment, and their acquisition by humans when exposed to the environment.

## Data Availability

All data produced in the present work are contained in the manuscript.

## Abbreviations

AJOL: African Journal Online
ARB: Antibiotic-resistant bacteria
GES: Guiana extended-spectrum beta-lactamase
IMP: Imipenemase
KPC: *Klebsiella pneumoniae* carbapenemase
MMAT: Mixed Methods Appraisal Tool
NDM: New Delhi metallo beta-lactamase
OXA: Oxacillinases
PRISMA: Preferred Reporting Items for Systematic Reviews and Meta-Analyses
PROSPERO: International prospective register of systematic reviews
VIM: Verona Integron encoding metallo beta-lactamase.

## Ethics approval and consent to participate

Not applicable.

## Consent for publication

Not applicable.

## Authors’ contributions

All the authors made substantial contributions to the study’s conceptualization and design, data acquisition, data analysis, and/or manuscript drafting.

AY, MS, BS, and LSBM participated in the conceptual design and development of the current study. OH, AY, OT, and HM assisted in the design and analysis of the search strategies. OH, tested and conducted searches, screened and reviewed the literature, and performed data extraction and spreadsheet production, with the assistance of HM, who reviewed and screened studies and performed data extraction. When a consensus could not be reached, the articles were reviewed by AY and OT. OH, HM, and OT shared in data analysis, while IIS constructed geographic mapping, AY created the R code used to analyze extracted data, and led in the development of analytic strategies and methods. All the authors participated in the initial quality assessment of the included studies. OH drafted the manuscript, which was reviewed and revised by AY, OT, SB, and SM. LSBM coordinated and directed the study and reviewed and revised the manuscript. All authors read and approved the final manuscript.

## Appendix A. Supplementary data

